# Environmental risk score as a predictor for 25-year symptom and functional trajectories in first episode psychosis

**DOI:** 10.64898/2026.05.14.26353217

**Authors:** Saheed Lawal, Jeffrey Liu, Olivia McLeron, Yuan Yang, Wenxuan Lian, Evangelos Vassos, Roman Kotov, Katherine Jonas

**Affiliations:** Department of Neurobiology and Behavior, Stony Brook University, NY 11794; Department of Psychiatry and Behavioral Health, Stony Brook University, NY 11794; Department of Psychology, Stony Brook University, NY 11794; Program in Public Health, Stony Brook University, NY,11794; Department of Applied Mathematics and Statistics, Stony Brook University, NY, 11794; Social, Genetic and Developmental Psychiatry (SGDP) Centre, Institute of Psychiatry, Psychology and Neuroscience, King’s College London, London, UK

**Keywords:** Environmental risk score, Schizophrenia, First Episode Psychosis, SCMHP, Psychosis Trajectories, Positive Symptoms

## Abstract

**Background and Hypothesis:** Environmental exposures linked to schizophrenia may play a role in shaping long-term clinical outcomes among individuals with psychotic disorders. This study examined whether the Maudsley Environmental Risk Score (ERS), a cumulative measure of five established environmental risk factors, predicts trajectories of symptoms, cognition, and psychosocial functioning over 25 years following first hospitalization for psychosis.

**Study Design:** Participants were drawn from the Suffolk County Mental Health Project, a longitudinal cohort of individuals with first-admission psychosis assessed six times over two decades. A total of 516 participants had sufficient ERS data and repeated assessments of symptoms (SAPS, SANS), cognitive ability, and functioning (GAF).

**Study Results:** Linear mixed-effects models showed that higher ERS was significantly associated with lower global functioning (β = -0.104, p = 0.008), an effect that remained consistent over time. ERS also predicted more severe and worsening reality distortion (β = 0.082, p = 0.023 for intercept; β = 0.005, p = 0.032 for slope of time). No significant associations were observed between ERS and cognitive ability, disorganization, or negative symptoms.

**Conclusions:** These findings highlight the contribution of environmental risk to functional impairment and persistent positive symptoms across the course of psychotic illness. Incorporating ERS into clinical risk models may aid the identification of individuals likely to experience a more severe illness trajectory, and inform long-term treatment planning.

## INTRODUCTION

Schizophrenia and psychotic disorders are associated with both genetic and environmental risk factors. Urbanicity, paternal age, minority status, cannabis use, childhood adversity, and obstetric complications each have a robust association with the incidence of schizophrenia ^1,2^. While each of these risk factors is independently associated with schizophrenia, measures of aggregate risk have the potential to improve power in studies of psychosis onset and course ^3^.

The Maudsley environmental risk score (ERS) is a weighted sum of established environmental risk factors for schizophrenia, with weights reflecting the strength of each factor’s association with the disorder. Overall, cumulative environmental risk is consistently elevated among individuals with psychotic disorders, hinting at the potential utility of ERS in research on illness onset and progression. Similar to polygenic risk scores, ERS are designed primarily as predictive tools rather than mechanistic explanations, summarizing multiple heterogeneous exposures into a single measure. As such, ERS capture aggregate risk across diverse social, behavioral, and environmental domains rather than representing a single underlying causal construct.

Associations between ERS and case-control status are relatively consistent. Mas et al. (2020) examined both ERS and polygenic risk scores (PRS) in a cohort of individuals with first-episode psychosis (FEP). They found that while the PRS alone was not predictive of case status, the ERS significantly differentiated cases from controls and explained a comparable proportion of variance as in other studies using similar cumulative environmental models. Moreover, Mas et al. reported a positive additive gene-environment interaction between high ERS and high PRS, indicating that individuals with both elevated environmental and genetic risk had disproportionately higher risk of psychosis, supporting the idea that environmental load and genetic liability act synergistically. These findings support the utility of cumulative environmental models and their interplay with genetic predisposition in psychosis risk^4^. Rami et al. (2025) assessed the influence of genetic and environmental risk scores on schizophrenia spectrum disorders in a case control cohort. Although their environmental score excluded cannabis use and migration status, they nonetheless observed an effect of ERS on case status ^5^. In a European case-control sample, Rodriguez et al. (2024) investigated the independent and joint effects of PRS and ERS on FEP. ERS included childhood adversity, cannabis use, migration, and paternal age. Both PRS and ERS independently predicted psychosis ^6^.

It is unknown whether ERS is associated with the longitudinal course of psychosis. Studies of population-based or at-risk populations have had mixed results. A longitudinal study of US youth found a higher baseline ERS predicted psychotic-like experiences and cognitive deficits 11 years later ^7^. Padmanabhan et al. (2017) assessed an ERS aggregating weighted environmental exposures in individuals at familial high risk for psychosis. They found that higher ERS significantly predicted transition to psychosis over follow-up ^8^. However, in a Brazilian adolescent cohort de Oliveira et al. (2021) examined whether ERS predicted psychotic experiences but found no significant associations. This may indicate sensitivity of ERS to age and culture ^9^. More specifically, because a majority of research on environmental risks for schizophrenia is based on data from the United Kingdom, Sweden, Denmark, and the United States, ERS may not reflect risks encountered in other parts of the world.^10^ Studies suggest that the increased risk of psychosis observed in some ethnic minority populations may partly reflect social and structural factors rather than ethnicity itself. In one analysis, social disadvantage and linguistic distance from the majority population were associated with increased odds of first-episode psychosis and partially mediated the association between minority status and psychosis risk ^11^. In addition, structural inequalities, migration-related experiences, and processes related to social exclusion have been proposed as mechanisms contributing to elevated psychosis risk in minority populations ^12^. Consistent with this perspective, recent evidence from low- and middle-income settings demonstrates that exposure to adverse social environments, such as childhood maltreatment occurring within contexts of economic precarity, disrupted caregiving systems, and limited social support, is strongly associated with psychotic outcomes, with cumulative exposure to multiple adversities showing particularly large effects ^13^. These findings support an ecological framework in which vulnerability is not inherent to minority status, but rather that broader structural conditions shape exposure to environmental risks among minority individuals.

Studies of the ERS as a predictor of course among cases have been inconclusive. Ferraro et al. (2025) estimated the Maudsley ERS in a large FEP sample and found that higher environmental score was associated with more severe cognitive decline prior to illness onset ^14^.In contrast, Cuesta et al. (2024) evaluated the prognostic value of an environmental risk score in a treated FEP sample and found no significant association between the ERS and key outcomes such as symptomatic remission, psychosocial functioning, or personal recovery ^15^. Collectively, studies indicate ERS predicts the course of cognition and psychotic-like symptoms up to the point of illness onset, but there is currently no evidence that ERS predicts symptom progression after onset.

In this study, we test whether the cumulative burden of environmental risk impacts symptoms and functional trajectories over the 25 years following first admission for psychosis. We hypothesized that higher ERS would predict a more severe course of positive symptoms, but also that the ERS might predict negative symptoms and cognition, which are core symptom dimensions of schizophrenia. If the proposed analyses support the above hypothesis, this suggests the potential of ERS, in combination with genetic risk, to be incorporated into clinical prediction models. The aim of such models would be to identify at first admission those individuals at risk for a more severe course of illness, who might benefit most from long-term, comprehensive care.

## METHOD

### Participants

Data were drawn from the Suffolk County Mental Health Project, a longitudinal first-admission study of psychosis. Between 1989 and 1995, individuals with first-admission psychosis were recruited from the 12 inpatient facilities in Suffolk County, New York (response rate 72%). The Stony Brook University Committee on Research Involving Human Subjects and the review boards of participating hospitals approved the protocol annually. Written consent was obtained from all study participants or their parents, for those who were minors at baseline. Eligibility criteria included residence in Suffolk County, age between 15 and 60, ability to speak English, absence of intellectual disability that would preclude consent and completion of the interview, first admission for psychosis within the past 6 months, and no apparent medical etiology for psychotic symptoms. A total of 516 participants met the inclusion criteria. Follow-up interviews were conducted at 6 months, 24 months, 48 months, 10 years, 20 years, and 25 years after baseline. Ninety-two participants died during the follow-up period.

The analysis sample included all participants with valid ERS data and at least two observations for any of the outcomes of interest: general cognitive ability, psychosocial function, or one of the four symptom domains. **Supplemental Table S1** compares the demographic and clinical characteristics of the analysis sample to those who were not included in the analysis because they did not meet these criteria for data completeness, for reasons other than mortality. Individuals not included in the analysis were four years older at baseline, on average, than those in the analysis. This likely reflects the difficulty of collecting information about premorbid risk factors from older individuals. There were no other clinical or demographic differences between the analysis sample and the full sample.

### Measures

Interviews were conducted by master level mental health professionals at each follow-up assessment. Medical records and interviews with individuals who know the participant well were also obtained at every assessment. This detailed information was used to complete the rating scales described below.

#### Diagnosis

Research diagnoses were made by consensus of study psychiatrists at baseline using all available information. The diagnostic process is outlined in Bromet and colleagues ^16^.

#### Age of Onset

Age of psychosis onset was determined based on symptom timelines obtained during first admission and 6-month follow-up diagnostic interviews conducted using the Structured Clinical Interview for DSM-III (SCID) at baseline and SCID-IV thereafter. This information was supplemented by informant interviews, school records, and medical records. For details see Jonas and colleagues ^17^.

#### Socioeconomic Status

Socioeconomic status was rated on the Hollingshead scale ^18^, which ranges from 1 (“large business owner/major professional/executive”) to 8 (“not working”). Ratings were based on the occupation of the primary earner in the participant’s household at baseline assessment.

#### Medication

Whether a participant was receiving antipsychotic medication at a given follow-up was coded as a binary variable. Data were derived from self-report, review of medication bottles brought to study appointments, review of medical records, and interviews with individuals who know the participants well.

#### Environmental Risk

Environmental risk factors were estimated using the procedure described in Vassos et al., 2020. Vassos included paternal age, cannabis use, childhood adversity, ethnic minority, urbanicity, and obstetric complications in their ERS. Assessment of these risks factors is described below.

<u>Paternal age</u> was assessed at baseline by self-report, and confirmed at the 6-month follow-up in interviews with participants’ parents and collaterals. <u>Cannabis use</u> was assessed at baseline using the Structured Clinical Interview for DSM-III-R ^19^. Cannabis use was operationalized as an ordinal variable which was 0 if the individual had no history of cannabis abuse or dependence, 1 if they had a lifetime history of cannabis use, and 3 if they had a lifetime history of cannabis abuse or dependence. <u>Childhood adversity</u> was based on information collected from the PTSD module of the Composite International Diagnostic Interview administered at the 24-month follow-up, as well as reports of lifetime history of stressful life events obtained during the 6-month follow-up interview ^20^. Childhood adversity was dichotomised such that participants reporting no exposure were assigned a score of -1.5, whereas any exposure to childhood adversity was assigned a score of 2.5 in accordance with the ERS scoring framework. A consensus procedure was used to review all material and determine age at exposure ^21^. <u>Minority status</u> was based on self-reported race and ethnicity collected at baseline. Ethnic minority status was included as part of the ERS as defined in prior work, and is interpreted here as a proxy for exposure to structural and social adversity rather than an individual-level biological risk factor. <u>Urbanicity</u> was assessed based on participants’ city or hospital of birth, with preference given to whichever location was more specific. Urbanicity was determined using either the participant’s reported city of birth or hospital of birth, with preference given to the more specific location; hospital of birth was used in 40% of cases. Maternity hospitals typically serve local geographic catchment areas and therefore generally reflect the residential environment at the time of birth. The median year of birth of individuals in the cohort was 1960, so population density of these locations was abstracted from 1960 census data^22,23^. <u>Obstetric complications</u> were assessed by parent report at the 6-month follow-up, but due to a high rate of missingness (64.9%), obstetric complications were excluded from this analysis. Of the remaining five risk factors, 30.4% of participants had complete data, 35.5% were missing data on one risk factor, and 28.8% were missing two risk factors. These participants were assigned a weight of 0 for the missing risk factors, meaning missing data does not upwardly or downwardly bias the ERS. Thirty-four participants (5.3%) were missing data on all but one or two risk factors. These individuals were removed from subsequent analyses.

The ERS for this study was calculated following the procedure established by Vassos *et al.* ^24^, with a slight adjustment adapted to reflect the prevalence and risk of psychosis in racial groups in the United States. The ERS was calculated according to the following steps (**Table 1**).

**Table 1:** Estimation of ERS from meta-analysis and scaled RR. ERS is composed of five factors that are subdivided into categories. Each sub-category is associated with a relative risk (RR) from meta-analysis. The RR is scaled using the population proportion (*p_j_*) of each sub-category. The logarithm of Scaled RR is used to estimate ERS for each sub-category. SCMHP = Suffolk County Mental Health Project.

| **Risk factor** | **Sub-categories** | **RR from meta-analysis** | **Population proportion ($p_i$)** | **Prevalence in SCMHP (p)** | **Scaled log(RR)** | **ERS** |
| --- | --- | --- | --- | --- | --- | --- |
| **Ethnic Minority** | American Indian | 1.62 | 0.011 | 0.005 | 0.16 | 1.5 |
|  | Black | 1.95 | 0.124 | 0.151 | 0.24 | 2.5 |
|  | White | 1.00 | 0.616 | 0.748 | -0.04 | -0.5 |
|  | Hispanic/Latino | 1.00 | 0.189 | 0.072 | -0.04 | -0.5 |
|  | Asian | 0.75 | 0.060 | 0.024 | -0.17 | -1.5 |
| **Paternal age** | < 40 | 1 | 0.921 | 0.877 | -0.01 | 0 |
|  | 40 - 50 | 1.17 | 0.071 | 0.111 | 0.06 | 0.5 |
|  | > 50 | 1.60 | 0.008 | 0.012 | 0.19 | 2 |
| **Cannabis use** | No Exposure | 1 | 0.700 | 0.588 | -0.12 | -1 |
|  | Abuse | 1.41 | 0.150 | 0.107 | 0.02 | 0 |
|  | Dependency | 2.77 | 0.150 | 0.305 | 0.32 | 3 |
| **Childhood adversity** | No Exposure | 1 | 0.730 | 0.425 | -0.17 | -1.5 |
|  | Any Exposure | 2.78 | 0.270 | 0.575 | 0.27 | 2.5 |
| **Urbanicity** | Low | 1.16 | 0.333 | 0.390 | -0.14 | -1.5 |
|  | Medium | 1.55 | 0.333 | 0.274 | -0.01 | 0 |
|  | High | 2.07 | 0.333 | 0.336 | 0.11 | 1 |

The risk weights for each factor are reported in **Table 1**. Childhood adversity, cannabis use, paternal age and urbanicity follow the same scoring system as outlined by Vassos et al. Among the environmental exposures included in the ERS, childhood adversity and cannabis use receive the highest weights, reflecting evidence linking these factors to substantially increased risk of psychotic disorders ^24^. Exposure to multiple forms of childhood adversity, including abuse, neglect, bullying, and interpersonal threat, is associated with substantially elevated odds of later psychotic disorder and demonstrates dose-response relationships with cumulative adversity ^13,25,26^. For childhood adversity, exposure to any form of adversity before the age of 16 was assigned a value of 2.5, while no exposure was assigned -1.5. Cannabis use has also been consistently associated with increased risk of psychotic outcomes, with evidence suggesting a dose-dependent relationship ^24,25^. For cannabis use, participants with cannabis abuse or dependence were assigned a value of 3, those who reported prior cannabis use were assigned a value of 0, while those with no exposure were assigned -1. Paternal age greater than 50 was assigned a score of 2, 40 to 50 was assigned 0.5, and less than 40 years was assigned 0. The distribution of population density was split into three tertiles, and participants were associated with low, medium, or high-density areas. Individuals from places with high densities were assigned a score of 1, medium densities were assigned 0, and low densities were assigned - 1.5. Ethnic minority was adjusted to better reflect the risk by minority population in the US (**Supplemental Method 1**). Scores were summed across risk factors to estimate the total ERS. The distribution of ERSs is depicted in **Figure 1**.

**Figure 1:**
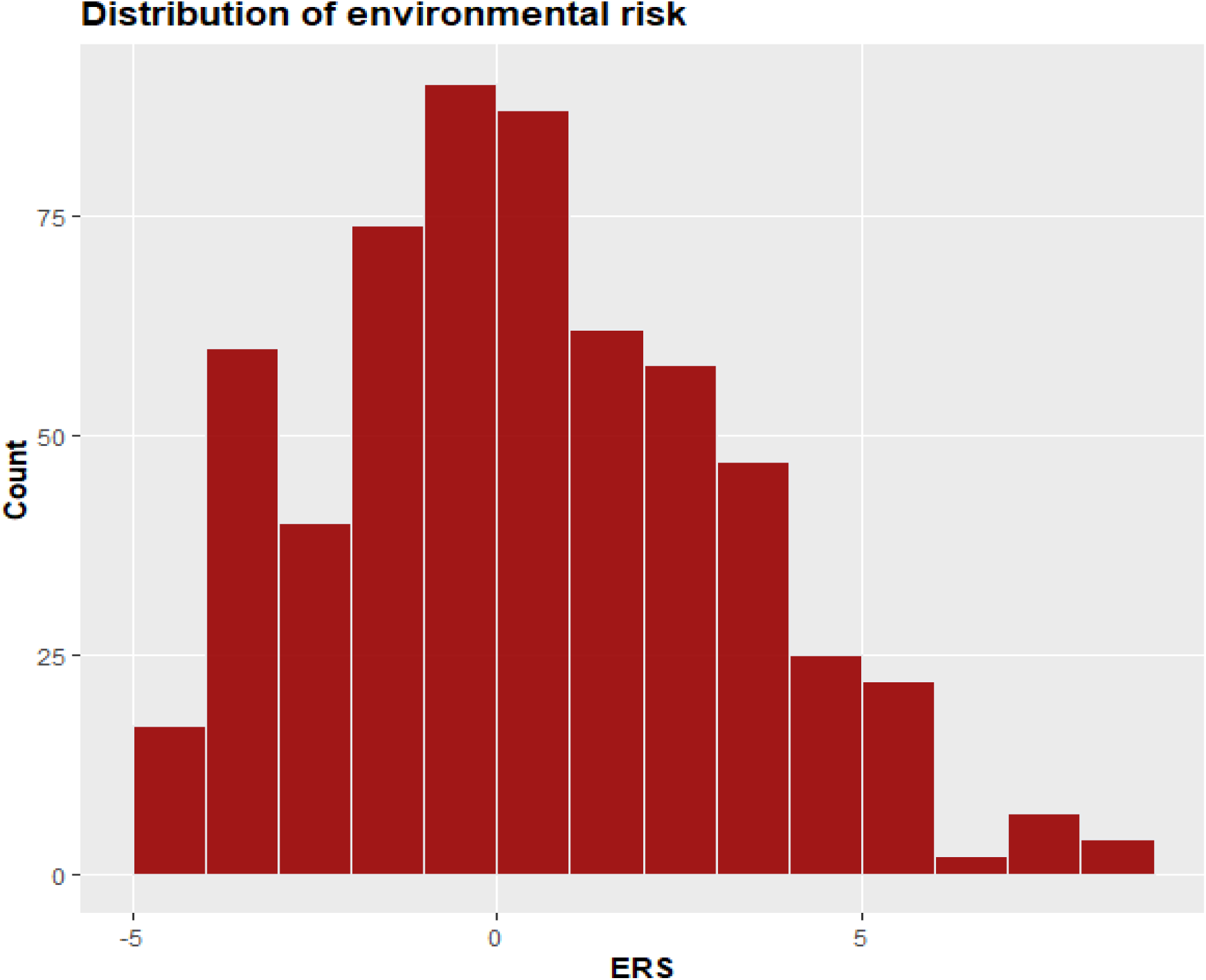
Distribution of ERS. ERS ranges from -4 to 9, with -0.5 and 1 as two most common scores. Mean ERS is 0.66 (SD = 2.77).

#### General Cognitive Ability

General cognitive ability was assessed at 6-month, 24-month, 20-year, and 25-year follow-ups. At 6- and 24-month follow-ups, cognitive ability was assessed using the Quick Test ^27^. The Quick Test is a brief measure of cognitive ability that is strongly correlated with scores on longer cognitive tests such as the WAIS ^28^. At the 24-month, 20-year, and 25-year follow-ups, general cognitive ability was estimated from results a neuropsychological battery including immediate trials of Verbal Paired Associates and Visual Reconstruction, Symbol-Digit Modalities, Trails A and B, the Controlled Oral Word Association Test, Vocabulary, and the Stroop Test. Parallel analysis indicated that a single factor accounted for the covariance among these test scores. A series of longitudinal structural equation models were used to establish configural, scalar, and metric invariance of the latent factor across follow-ups. Fit statistics of these models are reported in **Supplemental Table S9**. Latent trait estimates were extracted from this model. Since both neuropsychological test scores and Quick Test scores were available at the 24-month follow-up, a cross-walking procedure was used to translate neuropsychological test scores to the IQ metric. Latent trait estimates from cognitive models were cross-walked to the IQ metric by multiplying by the ratio of scores’ standard deviations, and then adding the difference in means. This process resulted in estimates of general cognitive ability at the 6-month, 24-month, 20-year, and 25-year follow-ups, harmonized to the IQ metric. See **Supplemental Method 2** and Jonas (2022) for additional detail.^29^

#### Psychosocial Function

Psychosocial function was rated using the Global Assessment of Function (GAF) scale. GAF is a numerical scale used to assess the severity of mental health symptoms and their impact on daily life. The GAF scale ranges from 1 to 90, with higher scores indicating better functioning and lower symptoms, while lower scores indicating more impairment and higher symptoms. GAF was rated at all major follow-up assessments (6-month, 24-month, 48-month, 10-year, 20-year, and 25-year) for the best month of the year preceding interview. Study psychiatrists used all available information (clinical interview, medical records, and collateral interviews) to generate consensus GAF ratings.

#### Symptom Severity

Symptom severity was assessed using the Scale for the Assessment of Positive Symptoms (SAPS) and Scale for the Assessment of Negative Symptoms (SANS). The SAPS and SANS comprise 49 items rated on a six-point scale (0 = none, 5 = severe). Ratings were made by interviewers using all available information at all major follow-up assessments, excluding baseline scores. Based on the results of structural analyses reported in Kotov et al (2016) ^30^, SAPS and SANS ratings were scored into four subscales: reality distortion, disorganization, inexpressivity, and avolition.

### Data analysis

ERS were estimated using Microsoft Excel and R ^31^. Contrasts between the analysis sample and its complement were performed using t-tests for continuous variables, and chi-square tests for categorical variables. Trajectories of symptoms, cognition, and functioning were estimated using mixed-effects longitudinal models based on all data available across follow-ups. Model fit was assessed using the Bayesian Information Criterion (BIC). Time was modeled relative to date of first admission and date of psychosis onset. For all phenotypes, modeling time relative to psychosis onset improved model fit (ΔBIC≈10-20 points), so all subsequent models were based on date of psychosis onset. Models included random intercepts and slopes for time, which for all phenotypes improved model fit (ΔBIC≥100 points). Adding a residual correlation structure did not improve model fit for a majority of phenotypes, so this parameter was not included in final models. The impact of environmental risk on cognitive, symptom, and functional trajectories was examined by including ERS as a fixed-effect predictor and by testing an ERS × time interaction term, which estimates whether ERS is associated with differential rates of change in outcomes over time. Code for all analyses is available at https://github.com/katherinegracejonas/ERS_psychosis_trajectories.

## RESULTS

### Demographic characteristics

The analysis sample includes 516 participants **(Table 2)**. The mean baseline age of the analysis sample was 29.6 years (SD = 9.4). Of the 516 participants, 294 were male (57.0%) and 222 were female (43.0%). Most participants identified as White (75.6%), followed by Black or African American (15.1%), Hispanic (7.2%), Asian or Pacific Islander (2.0%), and Native American (0.2%). Approximately 29.5% of the sample had a diagnosis of schizophrenia or schizoaffective disorder, followed by bipolar disorder with psychosis (23.1%), major depression with psychosis (16.1%), other psychosis (27.5%), and substance-induced psychosis (3.9%). Information about participants not included in the study is presented in **Supplemental Table S1.** We tested whether demographic and clinical variables were associated with missingness in environmental risk score components using one-way ANOVA (race, diagnosis), t-tests (sex), and correlation analyses (age, socioeconomic factor, and measures). Age was significantly associated with missingness (p = 0.026), whereas no other variables showed significant associations (all other p > 0.05) **(Supplemental Table S2)**

**Table 2:** Demographic characteristics: Participants demographics featuring mean age and socioeconomic status, racial/ethnic distribution, diagnostic breakdown, and baseline values for measures. N = total number with percentage, M = mean, and SD = standard deviation. Socioeconomic status is measured using Hollingshead’s rating of occupational prestige, which ranges from 1 to 8. A score of 1 indicates individuals who are not employed, 4 indicates skilled manual labor, and 8 indicates a business owner or person in a major professional role.

|  |  | **N (%)** | **M (SD)** |
| --- | --- | --- | --- |
|  | **Age at baseline** |  | 29.6 (9.4) |
|  | **Age at psychosis onset** |  | 27.5 (9.3) |
|  | **Socioeconomic status** |  | 5.4 (2.0) |
| **Sex** | **Male** | 294 (57.0) |  |
|  | **Female** | 222 (43.0) |  |
| **Race/Ethnicity** | **Black, not of Hispanic origin** | 78 (15.1) |  |
|  | **Hispanic** | 37 (7.2) |  |
|  | **White, not of Hispanic origin** | 390 (75.6) |  |
|  | **American Indian or Alaska native** | 1 (0.2) |  |
|  | **Asian or Pacific Islander** | 10 (1.9) |  |
| **Diagnosis** | **Schizophrenia / schizoaffective disorder** | 152 (29.5) |  |
|  | **Bipolar disorder with psychosis** | 119 (23.1) |  |
|  | **Major depression with psychosis** | 83 (16.1) |  |
|  | **Other psychosis** | 142 (27.5) |  |
|  | **Substance induced psychosis** | 20 (3.9) |  |
| **Measures at Baseline** | **Disorganization** |  | 5.1 (5.8) |
|  | **Reality Distortion** |  | 9.4 (8.4) |
|  | **Inexpressivity** |  | 7.0 (7.8) |
|  | **Avolition/Apathy** |  | 8.6 (6.7) |
|  | **GAF** |  | 58.8 (14.3) |
|  | **6 month IQ** |  | 97.6 (13.7) |

### Association between ERS and trajectories

The results of mixed effects models evaluating the association between ERS and trajectories of general cognitive ability, psychosocial function, and symptom severity are reported in **Table 3**. Higher ERS was significantly associated with lower GAF scores (β = -0.104, 95% CI = [-0.182, - 0.027], p = 0.008), although the ERS x time interaction was not significant (β = 0.001, 95% CI = [-0.003, 0.005], p = 0.722), indicating that ERS was associated with overall differences in global functioning but not with differential rates of change in functioning over time. ERS was not significantly associated with overall differences in general cognitive ability (β = -0.009, 95% CI = [-0.097, 0.079], p = 0.843), nor with cognitive trajectories (β = 0.004, 95% CI = [0.000, 0.008], p = 0.080).

**Table 3:** Interaction between ERS and measures: Mixed effect models showing the unstandardized (B) and standardized (β) values of the association between ERS and different measures over time (relative to symptom onset). Measures or phenotypes include positive symptoms (disorganization and reality distortion), negative symptoms (inexpressivity and avolition/apathy), GAF, and IQ. CI = confidence interval (95%). p-values less than 0.05 are considered significant. ^†^Only ERS and outcomes were standardized, not time, meaning coefficients reflect standardized rate of change in outcomes per year.

| | N | Parameter | B | 95% CI | $\beta^{\dagger}$ | 95% CI | p |
| --- | --- | --- | --- | --- | --- | --- | --- |
| **Disorganization (SAPS D)** | 479 | Intercept | 1.362 | [1.060, 1.663] | -0.339 | [-0.398, -0.280] | 0.000 |
|  |  | Time | 0.146 | [0.121, 0.171] | 0.030 | [0.025, 0.035] | 0.000 |
|  |  | ERS | 0.027 | [-0.077, 0.130] | 0.015 | [-0.043, 0.073] | 0.614 |
|  |  | ERS x time | 0.002 | [-0.006, 0.011] | 0.001 | [-0.004, 0.006] | 0.602 |
| **Reality distortion (SAPS P)** | 479 | Intercept | 2.065 | [1.636, 2.495] | -0.109 | [-0.179, -0.038] | 0.003 |
|  |  | Time | 0.057 | [0.027, 0.087] | 0.011 | [0.006, 0.016] | 0.000 |
|  |  | ERS | 0.172 | [0.024, 0.320] | 0.082 | [0.011, 0.152] | 0.023 |
|  |  | ERS x time | 0.011 | [0.001, 0.022] | 0.005 | [0.000, 0.010] | 0.032 |
| **Inexpressivity (SANS E)** | 452 | Intercept | 5.442 | [4.778, 6.107] | -0.072 | [-0.154, 0.010] | 0.084 |
|  |  | Time | 0.050 | [0.009, 0.091] | 0.006 | [0.001, 0.011] | 0.012 |
|  |  | ERS | -0.058 | [-0.286, 0.169] | -0.021 | [-0.102, 0.060] | 0.615 |
|  |  | ERS x time | 0.001 | [-0.013, 0.015] | 0.000 | [-0.005, 0.005] | 0.871 |
| **Avolition/Apathy (SANS A)** | 479 | Intercept | 6.523 | [5.953, 7.093] | -0.330 | [-0.402, -0.258] | 0.000 |
|  |  | Time | 0.224 | [0.190, 0.258] | 0.029 | [0.025, 0.034] | 0.000 |
|  |  | ERS | 0.176 | [-0.020, 0.372] | 0.064 | [-0.007, 0.136] | 0.080 |
|  |  | ERS x time | 0.003 | [-0.009, 0.015] | 0.001 | [-0.003, 0.005] | 0.611 |
| **GAF** | 516 | Intercept | 63.162 | [61.818, 64.506] | 0.536 | [0.459, 0.613] | 0.000 |
|  |  | Time | -0.699 | [-0.767, -0.631] | -0.041 | [-0.045, -0.037] | 0.000 |
|  |  | ERS | -0.636 | [-1.106, -0.166] | -0.104 | [-0.182, -0.027] | 0.008 |
|  |  | ERS x time | 0.004 | [-0.019, 0.028] | 0.001 | [-0.003, 0.005] | 0.722 |
| **IQ** | 392 | Intercept | 97.225 | [95.951, 98.499] | 0.262 | [0.173, 0.351] | 0.000 |
|  |  | Time | -0.317 | [-0.374, -0.260] | -0.022 | [-0.026, -0.018] | 0.000 |
|  |  | ERS | -0.044 | [-0.476, 0.388] | -0.009 | [-0.097, 0.079] | 0.843 |
|  |  | ERS x time | 0.017 | [-0.002, 0.037] | 0.004 | [0.000, 0.008] | 0.080 |

The main effect of ERS on reality distortion was significant (β = 0.082, 95% CI = [0.011, 0.152], p = 0.023). The interaction of ERS x time was also significant, indicating higher ERSs were associated with worsening hallucinations and delusions (β = 0.005, 95% CI = 0.000, 0.010], p = 0.032). **Figure 2** depicts the model-implied trajectories of reality distortion for hypothetical individuals with an ERS in the 25%ile versus 75%ile. Both individuals experienced worsening psychosis over 25 years, but individuals in the 75%ile experienced a more severe course. There was no significant association between ERSs and disorganization (β = 0.015, 95% CI = [-0.043, 0.073], p = 0.614), nor was there a significant interaction between ERS and time (β = 0.001, 95% CI = [0.004, 0.006], p = 0.602). To address potential confounding by cannabis use, we conducted a sensitivity analysis covarying for cannabis use status at each follow-up in our model. The association between high ERS and severity of positive symptoms remain significant after controlling for cannabis use, indicating that the observed effect is not attributable to continued cannabis consumption (**Supplemental Table S3**). The effects remained significant after controlling for antipsychotic medications as well **(Supplemental Table S4)**. These effects were not driven by minority status, as removal of this risk factor resulted in parameter estimates of a similar direction and magnitude **(Supplemental Table S5)**. The effect of ERS on reality distortion remains significant after covarying for sex, socioeconomic status, and age, but the effect on global functioning did not **(Supplemental Table S6)**.

**Figure 2:**
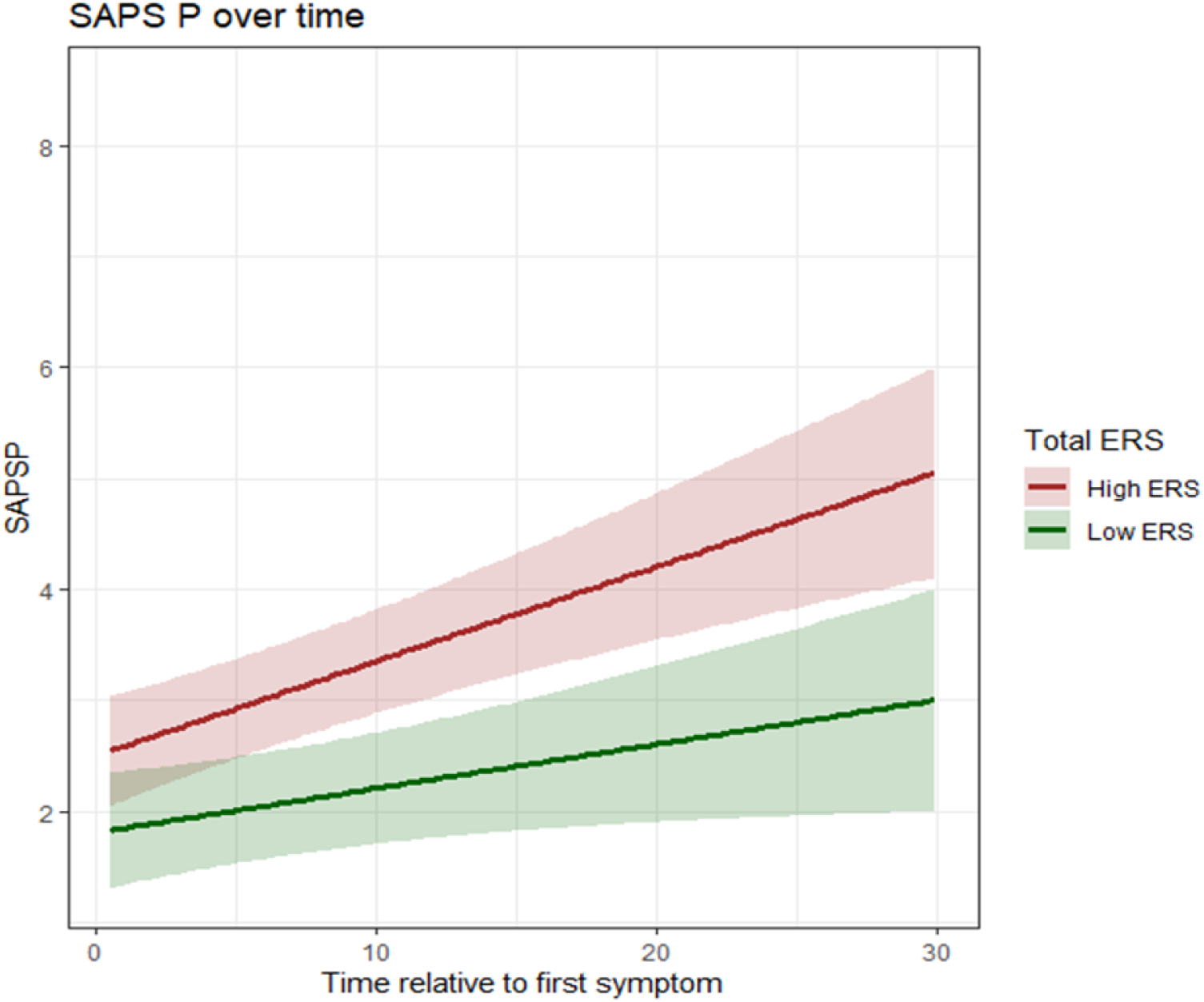
Effect of ERS on reality distortion: Model-implied trajectories of reality distortion (SAPS P) among participants with high vs low ERS over time. Individuals with high ERS (red line) exhibit a significantly higher rate of reality distortion progression over time compared to those with low ERS (green line) (β = 0.005, 95% CI = 0.000, 0.010], p = 0.032). NB: baseline symptoms not included.

There was no association between ERS and avolition/apathy (β = 0.064, 95% CI = [-0.007, 0.136], p = 0.080), and the ERS interaction with time was not significant (β = 0.001, 95% CI = [-0.003, 0.005], p = 0.611). Inexpressivity, the other measure of negative symptoms, also showed no significant association with ERS (β = -0.021, 95% CI = [-0.102, 0.060], p = 0.615), nor any significant interaction with time (β = 0.000, 95% CI = [-0.005, 0.005, p = 0.871).

To evaluate the potential impact of missing ERS data, we conducted two sensitivity analyses. First, analyses restricted to participants with complete ERS data resulted in reduced sample size and increased standard errors; although statistical significance was attenuated, parameter estimates were consistent with the primary analyses **(Supplemental Table S7)**. Second, to examine an upper-bound scenario, missing ERS items were imputed with the maximum risk value. Under this assumption, the main effect of ERS on avolition/apathy and the ERS × time interaction on IQ reached statistical significance **(Supplemental Table S8)**. If missingness is associated with higher underlying risk, the true effects may lie closer to these upper-bound estimates.

## DISCUSSION

Numerous environmental factors increase risk for psychotic disorders. Previous research has demonstrated that an index of cumulative environmental risk is associated with case-control status, psychotic-like experiences, cognitive deficits, and transition to psychosis. These findings indicate a consistent link between environmental risk and the development of psychotic symptoms and cognition up to the point of illness onset, but whether environmental risk continues to impact trajectories thereafter remains unknown. This study investigated whether a cumulative ERS is associated with the course of psychotic disorders after illness onset.

In a sample of 516 individuals recruited at first admission for psychosis, the ERS predicted the severity and trajectory of symptoms and functioning over the following 25 years. More environmental risk was consistently associated with poorer global functioning and more severe hallucinations and delusions across the illness course. That the association between ERS and global functioning did not vary over time suggests that environmental risk may contribute to an enduring vulnerability that shapes long-term functional outcomes. This is consistent with recent work in environmental risk score research for schizophrenia, where data derived from the HAMLETT study showed that higher cumulative exposure has been linked to poorer functioning even after accounting for clinical and sociodemographic factors ^32^. The extent to which functional deficits precede psychosis onset, or persist regardless of the course of psychosis, is a subject worth further study.

Higher ERSs also predicted worsening hallucinations and delusions over time. Notably, the ERS was not associated with general cognitive ability, disorganization, avolition/apathy, or inexpressivity. In sum, environmental risk not only increases the risk of developing a psychotic disorder, but could have a lasting effect that adversely impacts the course of psychosis after illness onset. The association between ERS and long-term trajectories of psychosis could also index an individual’s likelihood of being exposed to a variety of environmental risks over the illness course. However, the persistence of ERS-positive symptom relationship after controlling for cannabis use suggests that this association reflects a baseline effect of ERS itself, independent of subsequent exposure to risk, at least in the case of cannabis.

The impact of cumulative environmental risk on illness course was specific to hallucinations and delusions, as well as functioning. Environmental risks are identified largely through case-control comparisons. Since the presence of positive symptoms is required for a diagnosis of psychotic disorders, while negative symptoms and cognitive deficits are not, ERS may be insensitive to these symptom dimensions. An alternative, non-mutually exclusive explanation is that environmental risk exerts its strongest effects during premorbid development, contributing to a latent vulnerability that is preferentially expressed in positive symptom domains and functional outcomes. Although negative symptoms and cognitive deficits can continue to change after illness onset, the lack of association with ERS suggests that these domains may be influenced by distinct mechanisms or risk pathways. Taken together, these findings may reflect both the way environmental risks are operationalized and genuine differences in how cumulative exposures relate to specific symptom dimensions. This aligns with the findings of Erzin *et al.,* 2023, which found individuals with greater environmental risk – which in that study encompassed childhood adversity, cannabis use, and winter birth – had worse functioning and a poorer course of positive symptoms over a 1-month follow-up period ^33^. Erzin and colleagues also observed an association between ERSs and the course of negative symptoms, which was not observed in these analyses. This might be due to differences in the risk factors included in the analyses. Their ERS does not include urbanicity or advanced paternal age, while minority status and obstetric complications are included as covariates.

The lack of associations between environmental risk and IQ, disorganization, avolition/apathy, and inexpressivity could reflect four factors. First, it may be that not all domains are equally shaped by environmental burden. Cannabis use, for example, has been consistently linked to positive symptoms, but links with negative symptoms are inconsistent ^34–36^. Second, cognition and negative symptoms may be shaped by different environmental risks than positive symptoms. A majority of research in this area has focused on risk for psychosis, which is mostly diagnosed based on the presence of positive symptoms, rather than negative symptoms and cognition. Equally potent risk factors for these domains may exist but be undiscovered ^25^. Third, environmental risk may impact the development of negative symptoms and cognition in the premorbid, but not post-onset, phase of illness. The lack of an association between ERS and post-onset course of negative symptoms and cognition would not negate the relevance of ERS to the premorbid development of these symptom domains. Data from clinical-high risk for psychosis cohorts may help clarify whether a window of sensitivity to environmental risk moderates associations with outcomes. Fourth, evidence from other studies of environmental risk score suggests that associations with negative symptoms may be observed under certain conditions. For instance, Jeon et al., 2022 reported cross-sectional associations between environmental risk burden and negative symptom severity, and Kromenacker et al. 2025 also identified associations with negative symptoms in analyses of clinical outcomes ^37,38^. Additionally, recent work by Setiani et al., 2025 found that the KorealZlPolyenvironmental Risk Score was significantly associated with negative symptom severity at baseline in a schizophrenia spectrum cohort, further indicating that environmental risk scores can relate to negative symptom levels in crosslZlsectional and shortlZlterm designs ^39^. Notably, these studies primarily examine symptom severity at baseline or over relatively short follow-up periods, whereas the present study focuses on long-term trajectories across the illness course. Differences in the length of follow-up, operationalization of outcomes, and cultural or geographic context may therefore contribute to variability in findings.

Notably, the observed effect sizes were relatively small. However, small effects are common in studies of complex psychiatric outcomes such as psychosis, where risk reflects the cumulative contribution of multiple environmental and genetic factors. In this context, modest associations may still be relevant when considered as part of broader risk prediction frameworks.

These results support the potential utility of ERS in both population and individual intervention. Reducing population exposures to these risks may serve as both primary and secondary prevention, reducing risk of illness onset but also improving outcomes among cases. When integrated with genetic risk and early clinical indicators, ERS may help identify individuals most likely to follow a more severe illness course, who may benefit from more intensive, long-term support. Such personalized models could enhance risk stratification and improve resource allocation in early psychosis services.

### Strengths and limitations

This study has several notable strengths. First, it leverages a uniquely rich, longitudinal dataset spanning 25 years following first admission for psychosis, allowing for the assessment of long-term symptom and functional trajectories. Second, it is the first study, to our knowledge, to demonstrate an association between a cumulative ERS and positive symptom severity and progression, highlighting a previously unexamined aspect of environmental contributions to illness course. Third, the ERS incorporates multiple empirically supported environmental exposures, enabling a more comprehensive estimate of cumulative risk than single-factor approaches. Finally, the use of mixed effects models accommodates repeated measures and individual-level variability, improving the robustness of the findings.

This study also has some limitations. The method for estimating environmental risk implies it is a measurable, discrete factor rather than a latent construct that sums over a diverse range of social, economic, cultural and behavioral influences. It further assumes environmental risks are (a) uncorrelated, (b) exchangeable, and (c) function additively rather than synergistically. While the ERS includes well-validated environmental risk factors, it may not capture the timing, intensity, or interaction of exposures over the life course. In addition, the ERS approach aggregates diverse environmental exposures into a single score and therefore assumes that individual factors contribute additively and independently, and that similar scores may arise from different combinations of exposures. In addition, factors in the ERS are imperfect measures of risk. Minority status is a relatively crude proxy for factors such as structural adversity and social exclusion, which mediate the association between minority status and psychosis risk, and urbanicity is likely a proxy for other measures of neighborhood disadvantage. ERS are primarily intended as predictive tools rather than mechanistic explanations of risk. Future research is needed to better understand how specific environmental factors interact and whether particular exposures may have greater relevance for prevention efforts. Moreover, the study sample is drawn from a single geographic region and may not be representative of populations across the United States or globally, limiting the generalizability of the findings. Furthermore, the handling of missing ERS components represents a limitation. When data on one or two environmental exposures were missing, these were coded as 0 to avoid inflating risk scores. However, if missingness was associated with higher environmental risk, ERS may have been underestimated for some participants.

## CONCLUSIONS

In conclusion, this study provides new evidence that cumulative environmental risk burden is associated with more severe and persistent symptoms, particularly positive symptoms, as well as poorer functional outcomes across the long course of psychotic illness. These findings highlight the enduring impact of baseline environmental exposures beyond illness onset and support the incorporation of ERS into integrative risk models to inform early intervention and individualized care strategies in psychosis. While ERS is predictive, it does not explain the mechanisms driving these outcomes. Future research should examine how ERS interacts with broader social factors such as access to treatment, culturally responsive care, discrimination, stigma, and trauma-informed interventions to clarify pathways toward more positive outcomes and refine early intervention strategies.

## Data Availability

Data are available from the corresponding author upon reasonable request.

## Acknowledgements

This work was supported by the National Institutes of Health (MH094398 and MH110434 to R.K.; R01MH135119 to R.K. and K.G.J; and the Scholars in Biomedical Sciences [T32GM148331] to SL). The authors thank Evelyn Bromet, founder of the cohort. They gratefully acknowledge the support of the participants and mental health community of Suffolk County for contributing their time and energy to this project. They are also indebted to dedicated efforts of study coordinators, interviewers for their careful assessments, and to the psychiatrists who derived the consensus diagnoses.

## Supplemental Method 1 - ERS for Ethnic Minority

Risk values for ethnic minority status were based on estimates of relative risk (RR) for nonaffective psychotic disorders reported in Chung *et al.* ^40^. RRs were scaled using the formula:

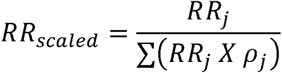

Where *p_j_* is the population prevalence of ethnic group *j*, and *RR_j_* is the relative risk of nonaffective psychosis in group *j*. The logarithm of RR_scaled_ was then multiplied by a constant of 10 and rounded up to the nearest half integer, in concordance with the procedure in Vassos et al. Risk weights were summed to estimate cumulative ERSs.

## Supplemental Method 2 - Conversion of test scores to IQ scale

All test scores were converted to the IQ scale (M=100; SD=15). Scores from the Quick Test were already on the IQ scale. Neuropsychological test scores at 24-month, 20-year, and 25-year follow-ups were converted to the IQ scale by first estimating a one-factor latent variable model from test scores, described in Jonas and colleagues (2019)^29,41^. Measurement invariance across time points was tested by first constraining loadings, then intercepts, and finally residuals to equality across time points. These constraints improved model fit **(Supplemental Table S9)**, indicating neuropsychological test scores are comparable across time points, and linking neuropsychological performance across time to a single scale. Both Quick Test scores and neuropsychological test scores were available at the 24-month assessment, which was used to translate latent factor scores to the IQ scale by multiplying latent scores by the ratio of Quick Test standard deviation to latent factor score standard deviation, then adding the difference in means.

## Tables

**Table S1:** Demographic characteristics of in-study vs out of study: Participants demographics featuring mean age and socioeconomic status, racial/ethnic distribution, diagnostic breakdown, and baseline values for measures. Participants in the analysis (N=516) are compared to those who attritted for reasons other than mortality (N=103). N = total number with percentage, M = mean, and SD = standard deviation.

|  |  | In Analysis<br>(N=516) |  | Out of Analysis<br>(Missing Outcome)<br>(N=70) |  | p | Out of Analysis (Missing<br>ERS)<br>(N = 33) |  |  |
| --- | --- | --- | --- | --- | --- | --- | --- | --- | --- |
|  |  | N (%) | M<br>(SD) | N (%) | M<br>(SD) |  | N (%) | M<br>(SD) | p |
|  | Age at baseline |  | 29.6<br>(9.4) |  | 33.9<br>(11.3) | 0.003 |  | 32.9<br>(9.6) | 0.046 |
|  | Age at psychosis<br>onset |  | 27.5<br>(9.3) |  | 30.7<br>(11.4) | 0.048 |  | 30.2<br>(10.2) | 0.463 |
|  | Socioeconomic<br>status |  | 5.4<br>(2.0) |  | 5.2<br>(2.2) | 0.589 |  | 5.7<br>(2.0) | 0.288 |
| Sex | Male | 294 (56.98) |  | 40 (57.1) |  | 1.000 | 23 (69.7) |  | 0.210 |
|  | Female | 222 (43.02) |  | 30 (42.9) |  |  | 10 (30.3) |  |  |
| Race/<br>Ethnicity | Black, not of<br>Hispanic origin | 78 (15.1) |  | 12 (17.1) |  | 0.885 | 5 (15.2) |  | 0.000 |
|  | Hispanic | 37 (7.2) |  | 3 (4.3) |  |  | 4 (12.1) |  |  |
|  | White, not of<br>Hispanic origin | 390 (75.6) |  | 54 (77.1) |  |  | 19 (57.6) |  |  |
|  | American Indian<br>or Alaska native | 1 (0.2) |  | 0 (0) |  |  | 1 (3.0) |  |  |
|  | Asian or Pacific<br>Islander | 10 (1.9) |  | 1 (1.4) |  |  | 5 (12.1) |  |  |
| Diagnosis | Schizophrenia /<br>schizoaffective<br>disorder | 152 (29.5) |  | 17 (24.3) |  | 0.179 | 12 (36.4) |  | 0.203 |
|  | Bipolar disorder<br>with psychosis | 119 (23.1) |  | 10 (14.3) |  |  | 2 (6.1) |  |  |
|  | Major depression<br>with psychosis | 83 (16.1) |  | 16 (22.9) |  |  | 8 (24.2) |  |  |
|  | Other psychosis | 142 (27.5) |  | 22 (31.4) |  |  | 10 (30.3) |  |  |
|  | Substance<br>induced<br>psychosis | 20 (3.9) |  | 5 (7.1) |  |  | 1 (3.0) |  |  |
| Measures<br>at<br>Baseline | Disorganization |  | 5.1<br>(5.8) |  | 4.8<br>(5.7) | 0.641 |  | 5.6<br>(5.4) | 0.654 |
|  | Reality Distortion |  | 9.4<br>(8.4) |  | 8.6<br>(9.5) | 0.484 |  | 8.5<br>(8.7) | 0.562 |
|  | Inexpressivity |  | 7.0<br>(7.8) |  | 6.2<br>(7.7) | 0.407 |  | 7.9<br>(8.1) | 0.516 |
|  | Avolition/Apathy |  | 8.6<br>(6.7) |  | 9.4<br>(7.1) | 0.335 |  | 10.1<br>(6.8) | 0.213 |
|  | GAF |  | 58.8<br>(14.3) |  | 57.0<br>(14.9) | 0.327 |  | 56.5<br>(14.7) | 0.372 |
|  | 6 month IQ |  | 97.6<br>(13.7) |  | 96.8<br>(13.3) | 0.766 |  | 93.9<br>(9.6) | 0.476 |

**Table S2:**
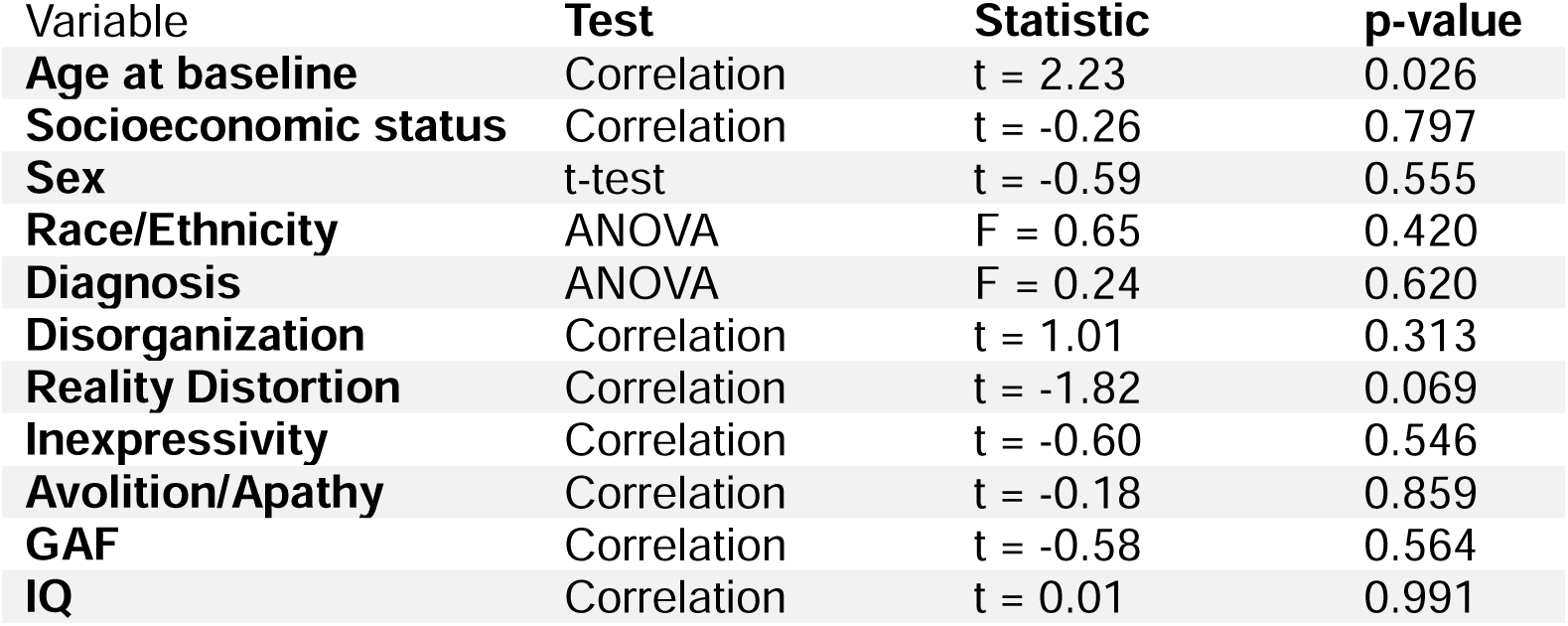
Association of demographics and measures with missingness in ERS.

| Variable | Test | Statistic | p-value |
| --- | --- | --- | --- |
| **Age at baseline** | Correlation | t = 2.23 | 0.026 |
| **Socioeconomic status** | Correlation | t = -0.26 | 0.797 |
| **Sex** | t-test | t = -0.59 | 0.555 |
| **Race/Ethnicity** | ANOVA | F = 0.65 | 0.420 |
| **Diagnosis** | ANOVA | F = 0.24 | 0.620 |
| **Disorganization** | Correlation | t = 1.01 | 0.313 |
| **Reality Distortion** | Correlation | t = -1.82 | 0.069 |
| **Inexpressivity** | Correlation | t = -0.60 | 0.546 |
| **Avolition/Apathy** | Correlation | t = -0.18 | 0.859 |
| **GAF** | Correlation | t = -0.58 | 0.564 |
| **IQ** | Correlation | t = 0.01 | 0.991 |

**Table S3:** Interaction between ERS and positive symptoms (covarying cannabis use): Mixed effect models showing the unstandardized (B) and standardized (β) values of the association between ERS and positive symptoms over time (relative to symptom onset), covarying for cannabis use. CI = confidence interval (95%). p-values less than 0.05 are considered significant.

| | N | Parameter | B | 95% CI | $\beta$ | 95% CI | p |
| --- | --- | --- | --- | --- | --- | --- | --- |
| **Disorganization (SAPS D)** | 479 | Intercept | 1.283 | [0.965, 1.600] | -0.357 | [-0.420, -0.294] | 0.000 |
|  |  | Time | 0.144 | [0.119, 0.170] | 0.030 | [0.025, 0.035] | 0.000 |
|  |  | ERS | 0.008 | [-0.097, 0.112] | 0.004 | [-0.055, 0.063] | 0.886 |
|  |  | ERS x time | 0.004 | [-0.005, 0.012] | 0.002 | [-0.003, 0.007] | 0.418 |
| **Reality distortion (SAPS P)** | 479 | Intercept | 1.745 | [1.296, 2.193] | -0.169 | [-0.243, -0.094] | 0.000 |
|  |  | Time | 0.065 | [0.035, 0.096] | 0.012 | [0.007, 0.017] | 0.000 |
|  |  | ERS | 0.124 | [-0.025, 0.273] | 0.059 | [-0.012, 0.129] | 0.105 |
|  |  | ERS x time | 0.011 | [0.000, 0.022] | 0.005 | [0.000, 0.010] | 0.042 |

**Table S4:** Interaction between ERS and positive symptoms (controlling for antipsychotics): Mixed effect models showing standardized (β) values of the association between ERS and positive symptoms over time (relative to symptom onset), after controlling for antipsychotic medications. CI = confidence interval (95%). p-values less than 0.05 are considered significant.

| | N | Parameter | $\beta$ | 95% CI | p |
| --- | --- | --- | --- | --- | --- |
| **Disorganization (SAPS D)** | 479 | Intercept | -0.328 | [-0.405, -0.250] | 0.000 |
|  |  | Time | 0.030 | [0.025, 0.035] | 0.000 |
|  |  | ERS | 0.005 | [-0.053, 0.062] | 0.878 |
|  |  | ERS x time | 0.002 | [-0.003, 0.007] | 0.487 |
| **Reality distortion (SAPS P)** | 479 | Intercept | -0.196 | [-0.288, -0.105] | 0.000 |
|  |  | Time | 0.012 | [0.007, 0.017] | 0.000 |
|  |  | ERS | 0.085 | [0.014, 0.156] | 0.019 |
|  |  | ERS x time | 0.006 | [0.001, 0.010] | 0.030 |

**Table S5:** Interaction between ERS and measures (without ethnic minority factor): Mixed effect models showing standardized (β) values of the association between ERS and measures over time (relative to symptom onset). CI = confidence interval (95%). p-values less than 0.05 are considered significant.

| | N | Parameter | $\beta$ | 95% CI | p |
| --- | --- | --- | --- | --- | --- |
| **Disorganization (SAPS D)** | 367 | Intercept | -0.343 | [-0.409, -0.277] | 0.000 |
|  |  | Time | 0.029 | [0.024, 0.035] | 0.000 |
|  |  | ERS | -0.035 | [-0.100, 0.031] | 0.299 |
|  |  | ERS x time | 0.003 | [-0.002, 0.009] | 0.197 |
| **Reality distortion (SAPS P)** | 367 | Intercept | -0.098 | [-0.180, -0.016] | 0.020 |
|  |  | Time | 0.010 | [0.004, 0.015] | 0.001 |
|  |  | ERS | 0.036 | [-0.046, 0.117] | 0.396 |
|  |  | ERS x time | 0.005 | [-0.001, 0.010] | 0.094 |
| **Inexpressivity (SANS E)** | 351 | Intercept | -0.089 | [-0.180, 0.002] | 0.056 |
|  |  | Time | 0.008 | [0.002, 0.013] | 0.008 |
|  |  | ERS | -0.076 | [-0.166, 0.015] | 0.103 |
|  |  | ERS x time | 0.003 | [-0.002, 0.009] | 0.269 |
| **Avolition/Apathy (SANS A)** | 367 | Intercept | -0.343 | [-0.424, -0.261] | 0.000 |
|  |  | Time | 0.030 | [0.025, 0.035] | 0.000 |
|  |  | ERS | 0.046 | [-0.036, 0.127] | 0.272 |
|  |  | ERS x time | 0.001 | [-0.004, 0.006] | 0.734 |
| **GAF** | 384 | Intercept | 0.548 | [0.459, 0.637] | 0.000 |
|  |  | Time | -0.041 | [-0.045, -0.037] | 0.000 |
|  |  | ERS | -0.065 | [-0.155, 0.025] | 0.156 |
|  |  | ERS x time | -0.001 | [-0.005, 0.003] | 0.702 |
| **IQ** | 318 | Intercept | 0.291 | [0.194, 0.387] | 0.000 |
|  |  | Time | -0.024 | [-0.028, -0.019] | 0.000 |
|  |  | ERS | 0.065 | [-0.032, 0.161] | 0.188 |
|  |  | ERS x time | 0.002 | [-0.002, 0.006] | 0.334 |

**Table S6:** Interaction between ERS and measures (covarying sex, socioeconomic factor and age at baseline): Mixed effect models showing standardized (β) values of the association between ERS and measures over time (relative to symptom onset), with sex, socioeconomic factor and age as covariates. CI = confidence interval (95%). p-values less than 0.05 are considered significant.

| | N | Parameter | $\beta$ | 95% CI | p |
| --- | --- | --- | --- | --- | --- |
| **Disorganization (SAPS D)** | 479 | Intercept | -0.295 | [-0.371, -0.219] | 0.000 |
|  |  | Time | 0.030 | [0.025, 0.035] | 0.000 |
|  |  | ERS | 0.002 | [-0.057, 0.061] | 0.942 |
|  |  | ERS x time | 0.001 | [-0.004, 0.006] | 0.616 |
| **Reality distortion (SAPS P)** | 479 | Intercept | -0.042 | [-0.132, 0.048] | 0.360 |
|  |  | Time | 0.011 | [0.006, 0.016] | 0.000 |
|  |  | ERS | 0.063 | [-0.008, 0.133] | 0.083 |
|  |  | ERS x time | 0.005 | [0.000, 0.010] | 0.033 |
| **Inexpressivity (SANS E)** | 452 | Intercept | 0.020 | [-0.081, 0.122] | 0.694 |
|  |  | Time | 0.006 | [0.001, 0.011] | 0.017 |
|  |  | ERS | -0.053 | [-0.132, 0.027] | 0.198 |
|  |  | ERS x time | 0.000 | [-0.005, 0.005] | 0.883 |
| **Avolition/Apathy (SANS A)** | 479 | Intercept | -0.178 | [-0.267, -0.090] | 0.000 |
|  |  | Time | 0.029 | [0.025, 0.033] | 0.000 |
|  |  | ERS | 0.036 | [-0.034, 0.105] | 0.314 |
|  |  | ERS x time | 0.001 | [-0.003, 0.005] | 0.638 |
| **GAF** | 516 | Intercept | 0.409 | [0.313, 0.505] | 0.000 |
|  |  | Time | -0.041 | [-0.045, -0.037] | 0.000 |
|  |  | ERS | -0.066 | [-0.142, 0.011] | 0.093 |
|  |  | ERS x time | 0.001 | [-0.003, 0.004] | 0.760 |
| **IQ** | 392 | Intercept | 0.253 | [0.146, 0.361] | 0.000 |
|  |  | Time | -0.021 | [-0.025, -0.017] | 0.000 |
|  |  | ERS | 0.026 | [-0.059, 0.111] | 0.547 |
|  |  | ERS x time | 0.004 | [0.000, 0.008] | 0.066 |

**Table S7:** Interaction between ERS and measures (Individuals with complete ERS data): Mixed effect models showing standardized (β) values of the association between ERS and measures over time (relative to symptom onset), using only data from individuals with complete ERS. CI = confidence interval (95%). p-values less than 0.05 are considered significant.

| | N | Parameter | $\beta$ | 95% CI | p |
| --- | --- | --- | --- | --- | --- |
| **Disorganization (SAPS D)** | 180 | Intercept | -0.368 | [-0.454, -0.282] | 0.000 |
|  |  | Time | 0.032 | [0.025, 0.040] | 0.000 |
|  |  | ERS | 0.024 | [-0.061, 0.109] | 0.573 |
|  |  | ERS x time | -0.001 | [-0.009, 0.007] | 0.816 |
| **Reality distortion (SAPS P)** | 180 | Intercept | -0.096 | [-0.202, 0.010] | 0.076 |
|  |  | Time | 0.009 | [0.000, 0.017] | 0.043 |
|  |  | ERS | 0.067 | [-0.039, 0.172] | 0.216 |
|  |  | ERS x time | 0.002 | [-0.007, 0.010] | 0.667 |
| **Inexpressivity (SANS E)** | 175 | Intercept | -0.104 | [-0.223, 0.015] | 0.089 |
|  |  | Time | 0.009 | [0.002, 0.016] | 0.016 |
|  |  | ERS | -0.084 | [-0.203, 0.034] | 0.165 |
|  |  | ERS x time | 0.007 | [0.000, 0.014] | 0.050 |
| **Avolition/Apathy (SANS A)** | 180 | Intercept | -0.340 | [-0.451, -0.229] | 0.000 |
|  |  | Time | 0.029 | [0.023, 0.036] | 0.000 |
|  |  | ERS | 0.043 | [-0.067, 0.153] | 0.445 |
|  |  | ERS x time | 0.008 | [0.001, 0.014] | 0.024 |
| **GAF** | 185 | Intercept | 0.564 | [0.440, 0.689] | 0.000 |
|  |  | Time | -0.042 | [-0.048, -0.036] | 0.000 |
|  |  | ERS | -0.114 | [-0.238, 0.011] | 0.076 |
|  |  | ERS x time | -0.001 | [-0.007, 0.005] | 0.792 |
| **IQ** | 165 | Intercept | 0.287 | [0.158, 0.417] | 0.000 |
|  |  | Time | -0.025 | [-0.031, -0.019] | 0.000 |
|  |  | ERS | 0.061 | [-0.068, 0.191] | 0.355 |
|  |  | ERS x time | 0.001 | [-0.005, 0.008] | 0.715 |

**Table S8:**
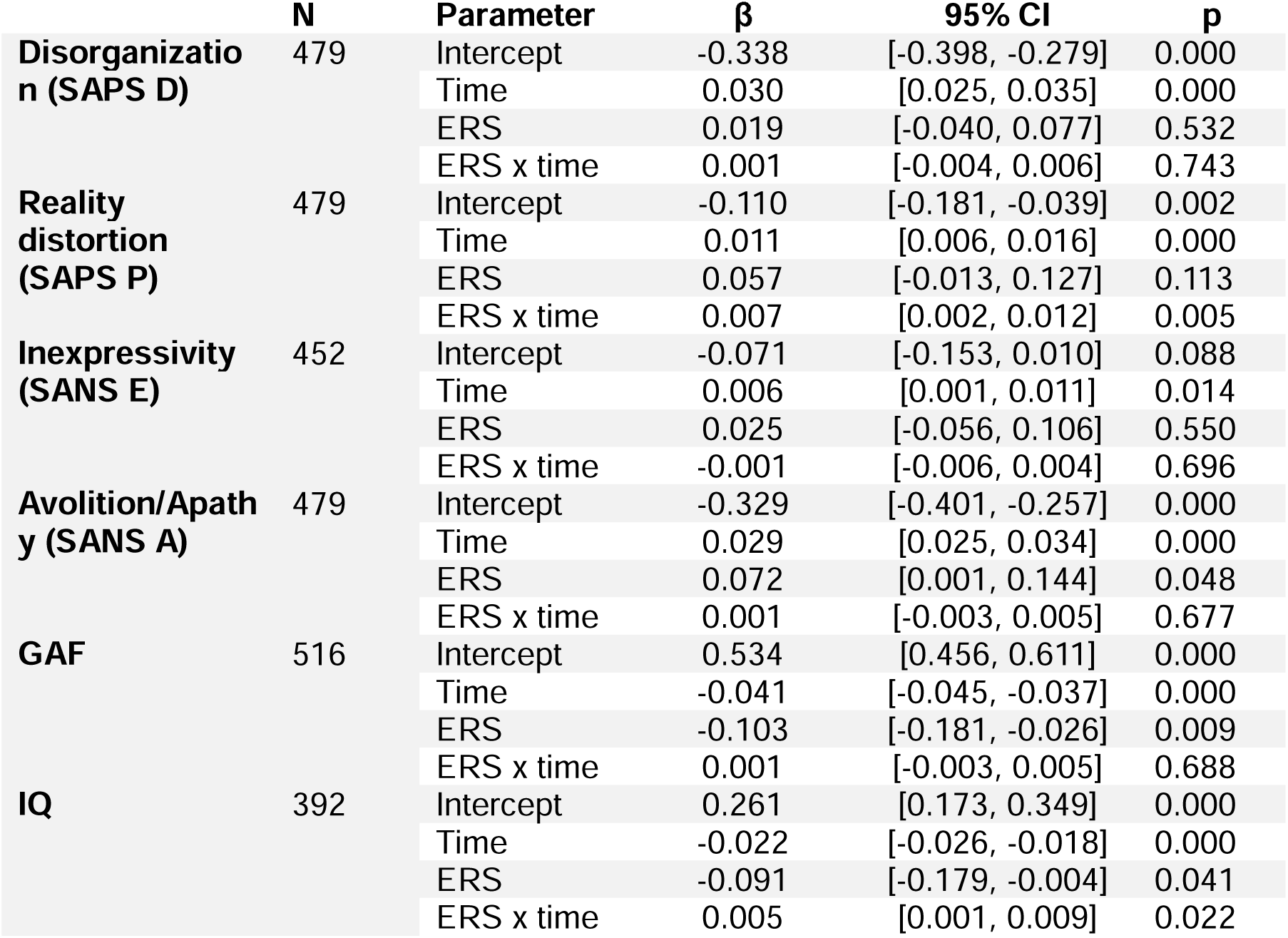
Interaction between ERS and measures (missing ERS imputed with the maximum risk value): Mixed effect models showing standardized (β) values of the association between ERS and measures over time (relative to symptom onset), with missing ERS imputed with the maximum risk value. CI = confidence interval (95%). p-values less than 0.05 are considered significant.

**Table S9:** Tests of measurement invariance for the neuropsychological test battery across follow-up assessments.

| Model | CFI | RMSEA | BIC |
| --- | --- | --- | --- |
| Equal structure | 0.746 | 0.097 | 48728.673 |
| Equal loadings | 0.770 | 0.090 | 48559.32 |
| Equal intercepts | 0.900 | 0.061 | 48066.055 |
| Equal residuals | 0.893 | 0.061 | 48024.154 |

